# Associations between metabolic heterogeneity of obesity and osteoarthritis: A prospective cohort study

**DOI:** 10.1101/2025.09.02.25334896

**Authors:** Hao Zhang, Hao Yang, Baiyong Zhu, Zhenghui Liao, Muhui Zeng, Jiawei Chen, Changhai Ding, David J Hunter, Zhaohua Zhu

**Author notes:** Correspondence: Z. Zhu. and C. Ding. Clinical Research Centre, Orthopedic Centre, Zhujiang Hospital, Southern Medical University, Guangzhou, China. Hao Zhang and Hao Yang contributed equally to this article.

## Abstract

**Background:** Previous studies have shown that obesity accelerates the development of osteoarthritis (OA). However, obesity is metabolically heterogeneous. The association between metabolic heterogeneity of obesity and incident OA remains unclear.

**Methods:** A total of 381,036 participants from the UK Biobank (UKBB) were included baseline. Metabolic heterogeneity of obesity was evaluated based on four obesity and metabolic phenotypes: metabolically healthy non-obesity (MHNO), metabolically unhealthy non-obesity (MUNO), metabolically healthy obesity (MHO), and metabolically unhealthy obesity (MUO). Incident OA cases were identified through self-reported diagnoses and hospital records. Multivariable-adjusted Cox proportional hazards models were used to evaluate the associations of these obesity phenotypes with OA incidence.

**Results:** In the UKBB (mean age 56.07 ± 8.13 years; 59.1% female; median follow-up 12.35 years [Interquartile range (IQR) 1.8 years]), the cohort included 246,565 MHNO, 30,960 MHO, 46,834 MUNO, and 56,677 MUO participants. Longitudinal analyses revealed distinct risk patterns between metabolic heterogeneity of obesity and OA development. For total OA, risk was elevated across all groups compared with MHNO: MUNO (HR 1.20, 95% CI 1.17–1.23), MHO (HR 1.72, 95% CI 1.68–1.77), and MUO (HR 1.87, 95% CI 1.83–1.91), with the highest risk observed in the MUO group, indicating a synergistic effect of obesity and metabolic dysfunction. This gradient pattern was particularly evident for knee OA, where MUO (HR 2.56, 95% CI 2.47, 2.66) had the greatest risk, followed by MHO (HR 2.42, 95% CI 2.31, 2.53) and MUNO (HR 1.23 [1.18, 1.29]). For hip OA, MUO (HR 1.49 [1.42, 1.56]) and MHO (HR 1.51 [1.42, 1.61]) showed similar elevations, while MUNO (HR 1.04 [0.99, 1.10]) were not significantly associated. For hand OA, MUO (HR 1.13 [1.02, 1.26]) had a moderate risk, slightly lower than MUNO (HR 1.18 [1.06, 1.31]), while MHO (HR 1.08 [0.94, 1.24]) showed no significant association. Importantly, metabolic dysfunction independently contributed to OA risk across all weight categories. Mediation analysis further indicated that metabolic factors explained approximately 15% of the BMI effect on total OA and 11% on knee OA, whereas BMI had no significant total effect on hand OA.

**Conclusions:** The study highlights the importance of maintaining and promoting metabolic health, particularly in overweight/obese individuals, to reduce OA risk. Metabolic factors were identified as key mediators of the association between BMI and OA in weight-bearing joints, emphasizing the need for targeted strategies addressing both metabolic dysfunction and obesity.

## Introduction

Osteoarthritis (OA) is a leading cause of pain, functional limitations, and disability(1). From 1990 to 2021, the global burden of OA rose markedly, now affecting over 595 million individuals, with metabolic dysfunction emerging as a key driver of regional disparities alongside genetic and behavioral factors(2). To date, no definitive therapy has been found to reverse the progression of OA or prevent cartilage degradation(3). Thus, managing obesity and improving metabolic health represent key strategies to mitigate OA risk in ageing populations.

The worldwide obesity epidemic is emerging as a major global health challenge(4). Previous studies indicated that obesity was associated with accelerated OA prevalence(5). However, these studies did not consider the metabolic heterogeneity of obesity. Epidemiological surveys indicated that approximately one-third of overweight and obese individuals were metabolically healthy, despite regional variations worldwide(6). Therefore, obesity has been further stratified into metabolically healthy obesity (MHO) and metabolically unhealthy obesity (MUO). Similarly, among individuals with non-obesity, metabolic heterogeneity is observed, allowing classification into metabolically healthy non-obesity (MHNO) and metabolically unhealthy non-obesity (MUNO)(7, 8). However, the impact of these distinct phenotypes on OA development has not been fully elucidated.

To address these gaps, we leveraged data from the UK Biobank (UKBB) prospective cohort to examine the associations between metabolic heterogeneity of obesity and incident OA, and to explore the interplay between adiposity and metabolic dysfunction in OA development.

## Methods

### Study populations

The UKBB is a population-based cohort study involving over 500,000 participants between the ages of 37 and 73. These individuals attended 1 of the 22 assessment centers located across the United Kingdom (UK) from 2006 to 2010(9). The study received approval from the Northwest Multi-center Research Ethics Committee (REC reference 11/NW/0382), and participants provided written informed consent before their involvement. The UKBB study was further approved by the National Health Service (NHS) National Research Ethics Service (16/NW/0274) for conducting sub-studies within it(9).

In the current study, a total of 381,036 participants were included in the main analysis after excluding participants who withdrew from the study (n = 1,298), those with prior diagnosis of OA at baseline (n = 55,940), and those with missing data on metabolic heterogeneity of obesity (n = 64,137) (Figure 1).

**Figure 1.**
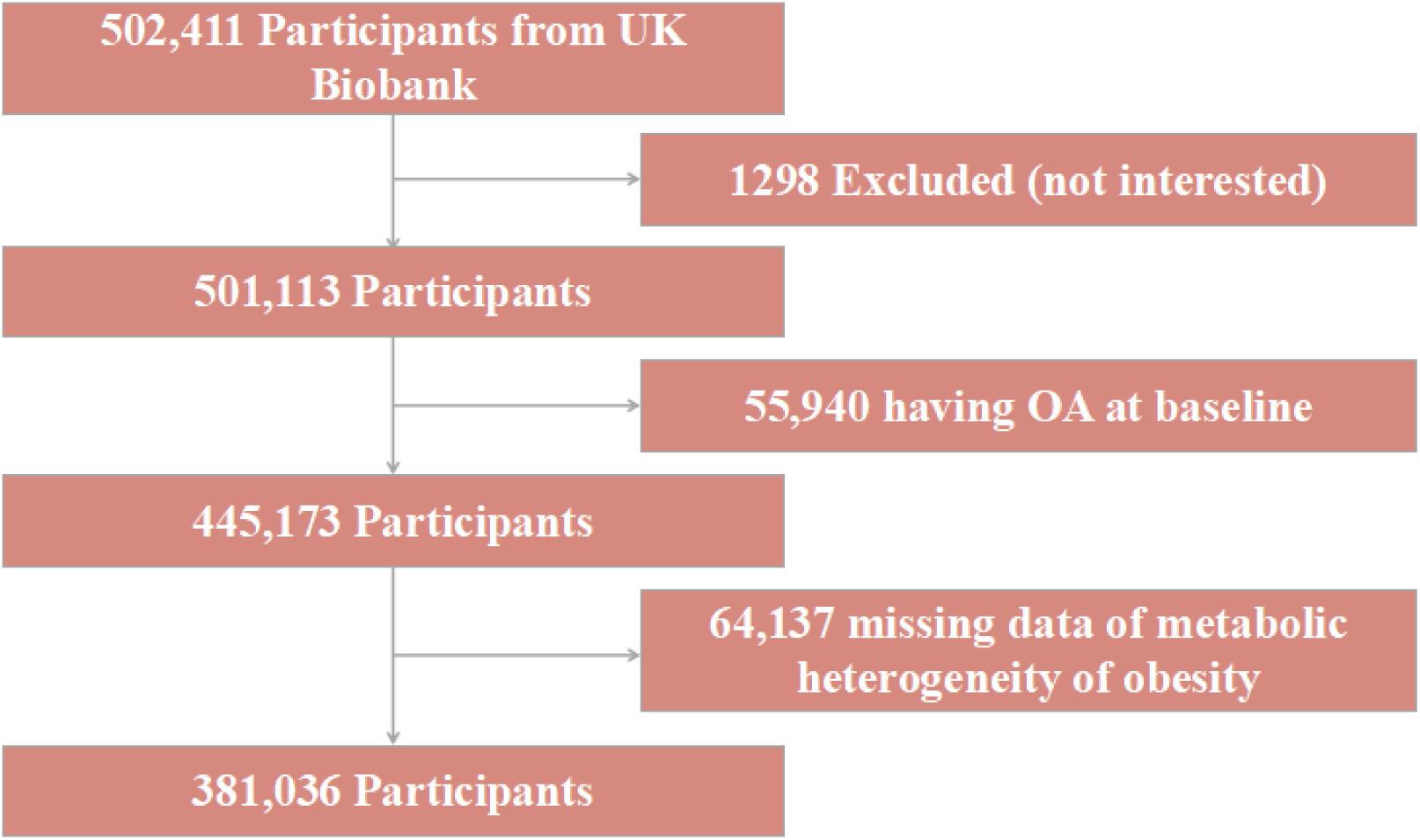
Flow chart of UK Biobank participants screening.

### Assessment of outcome

In UKBB, OA presence was identified through hospital inpatient records linked to Hospital Episode Statistics (HES) for England, the Scottish Morbidity Record, and the Patient Episode Database for Wales(10). Diagnoses were identified using ICD-9 (7151, 7152, 7153, 7158, 7159) and ICD-10 (M15–M19) codes, along with self-reported data (Field 20002)(9) (Supplementary Table 1).

### Assessment of metabolic heterogeneity of obesity

Obesity status was assessed by body mass index (BMI) based on country-specific criteria. Participants were categorized into two groups as non-obesity (18.5 ≤ BMI < 30.0 kg/m2) and obesity (BMI ≥ 30.0 kg/m2)(11). Metabolic status was assessed based on five metabolic components(9, 12). Participants who met three or more of the following five criteria were classified as metabolically unhealthy: abdominal obesity (waist circumference >88 cm in females or >102 cm in males), elevated blood pressure (systolic ≥135 mmHg or diastolic ≥85 mmHg) or antihypertensive medication use, fasting glucose ≥6.11 mmol/L or diabetes diagnosis/antidiabetic treatment, reduced HDL-C levels (<1.3 mmol/L in females or <1.04 mmol/L in males) or lipid-lowering therapy, and elevated triglycerides (≥1.70 mmol/L) or lipid-modifying drug use. This operational definition aligns with NCEP-ATPIII criteria, with thresholds calibrated to match standardized biochemical measurements in population-based biobank studies(13) (Supplementary Table 2). Combined with obesity and metabolic status, participants were divided into four BMI-metabolic phenotypes as MHNO, MUNO, MHO, and MUO to evaluate the metabolic heterogeneity of obesity.

### Assessment of covariates

Evidence from large-scale epidemiological studies indicates that OA could be associated with socioeconomic status and unhealthy lifestyle factors. According to these findings and a priori knowledge about our data, a set of covariates was ascertained and collected at baseline, including age, sex, race, education, Townsend deprivation index, healthy diet, smoking status, drinking status, metabolic equivalent task-physical activity (MET-PA), history of joint injury, and glucosamine use(14).

### Statistical analysis

For descriptive analyses, continuous variables are presented as mean (standard deviation [SD]) and categorical variables as counts (percentages). Baseline characteristics were compared using one-way ANOVA or Kruskal–Wallis rank sum tests for continuous variables and Chi-square tests for categorical variables.

To examine the associations between metabolic heterogeneity of obesity and incident OA, multivariable Cox regression models were fitted to estimate hazard ratios (HRs) with 95% confidence intervals (CIs), using the MHNO group as the reference. Two models were specified for the UKBB analyses: Model 1 was unadjusted, while Model 2 was adjusted for race, education, Townsend deprivation index, healthy diet, smoking status, drinking status, MET-PA, history of joint injury, and glucosamine use. The cumulative risk and survival probability of incident OA across groups were estimated using Kaplan–Meier (KM) curves with log-rank tests. Subgroup and interaction analyses were conducted by stratifying participants by age (<60 or ≥60 years) and sex (male or female). KM curves were also applied to explore the associations between metabolic heterogeneity of obesity and the cumulative risk of incident OA within these subgroups.

A causal mediation analysis was conducted to investigate whether metabolic factors mediate the relationship between BMI and incident OA. Cox regression models were fitted to estimate the associations between BMI, metabolic factors, and OA outcomes, adjusting for all the confounders included in Model 2. Specifically, the model provided estimates for the direct effect (DE), indirect effect (IE), total effect (TE), and the proportion mediated (PM)(15). The 95% CIs for these estimates were calculated using Bootstrap with 1,000 repetitions. The causal mediation analysis was conducted using R package CMAverse(16).

In the sensitivity analyses, we excluded individuals diagnosed with OA within the first and first two years of follow-up to mitigate the potential impact of reverse causality(17). All *P*-values were two-tailed, with significance defined as *P* < 0.05. Analyses and visualization were conducted using R software (version 4.2.1).

## Results

### Baseline characteristics

A total of 381,036 participants from UKBB were included after applying the exclusion criteria (Figure 1). Table 1 presents the baseline characteristics of the study population by obesity and metabolic status. The mean age of the participants was 56.07 ± 8.13 years, and 59.1% were female. The MUO phenotype exhibited the most adverse metabolic profile, characterized by the highest mean BMI (34.30 ± 3.97), SBP, glucose levels, triglyceride levels, and LDL concentrations, alongside the lowest HDL concentrations, with all *P*-values <0.001 compared to other groups. Socioeconomic disparities were evident in the Townsend deprivation indices, which increased from -1.14 in the MHNO group to -0.35 in the MUO group. Similarly, educational attainment showed significant variation, with 75.3% of MUO participants having lower education levels compared to 62.4% in the MHNO group. 48.5% of MUO individuals engaged in regular physical activity compared to 61.7% in the MHNO group (Table 1).

**Table 1.**
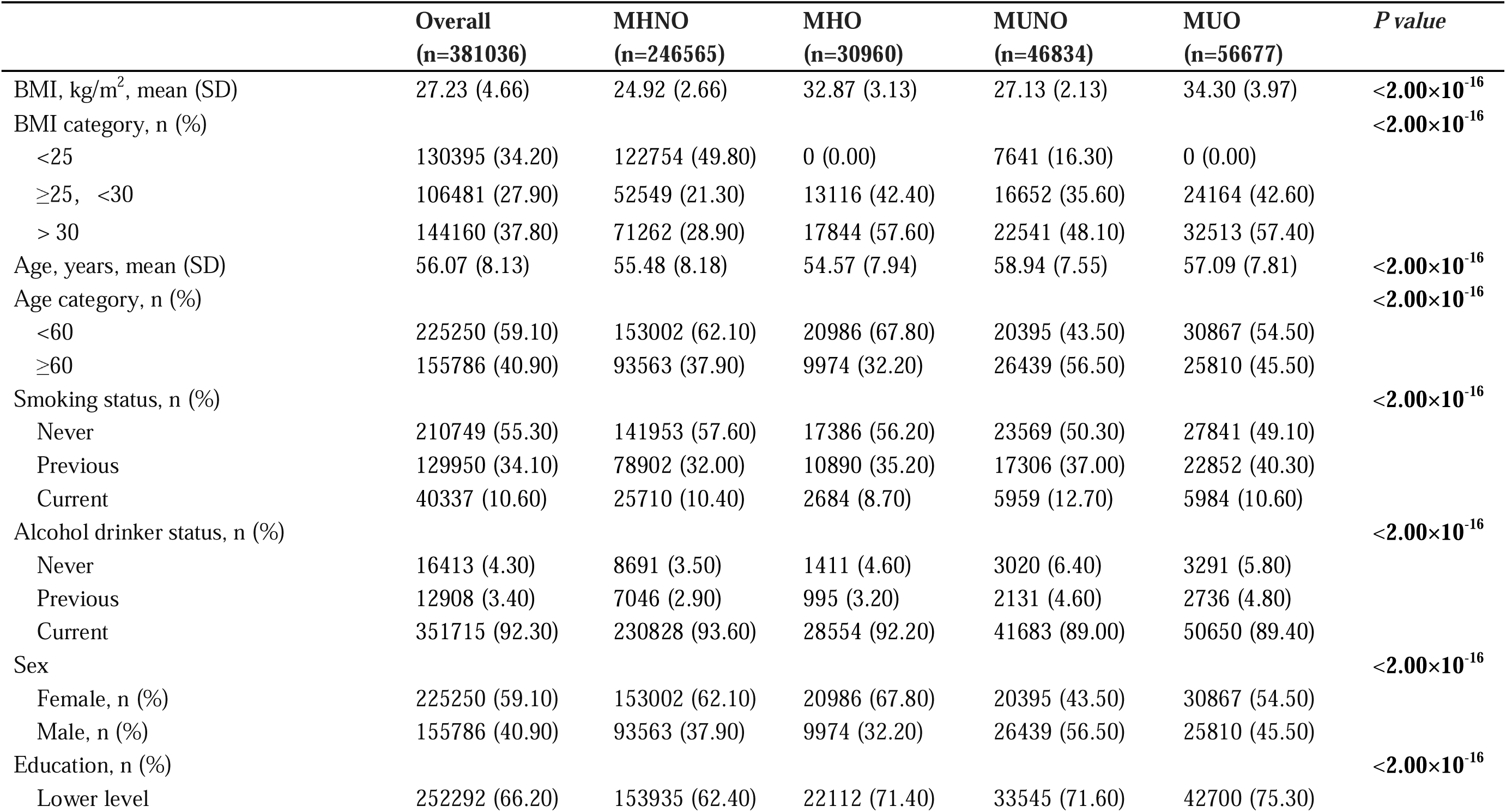
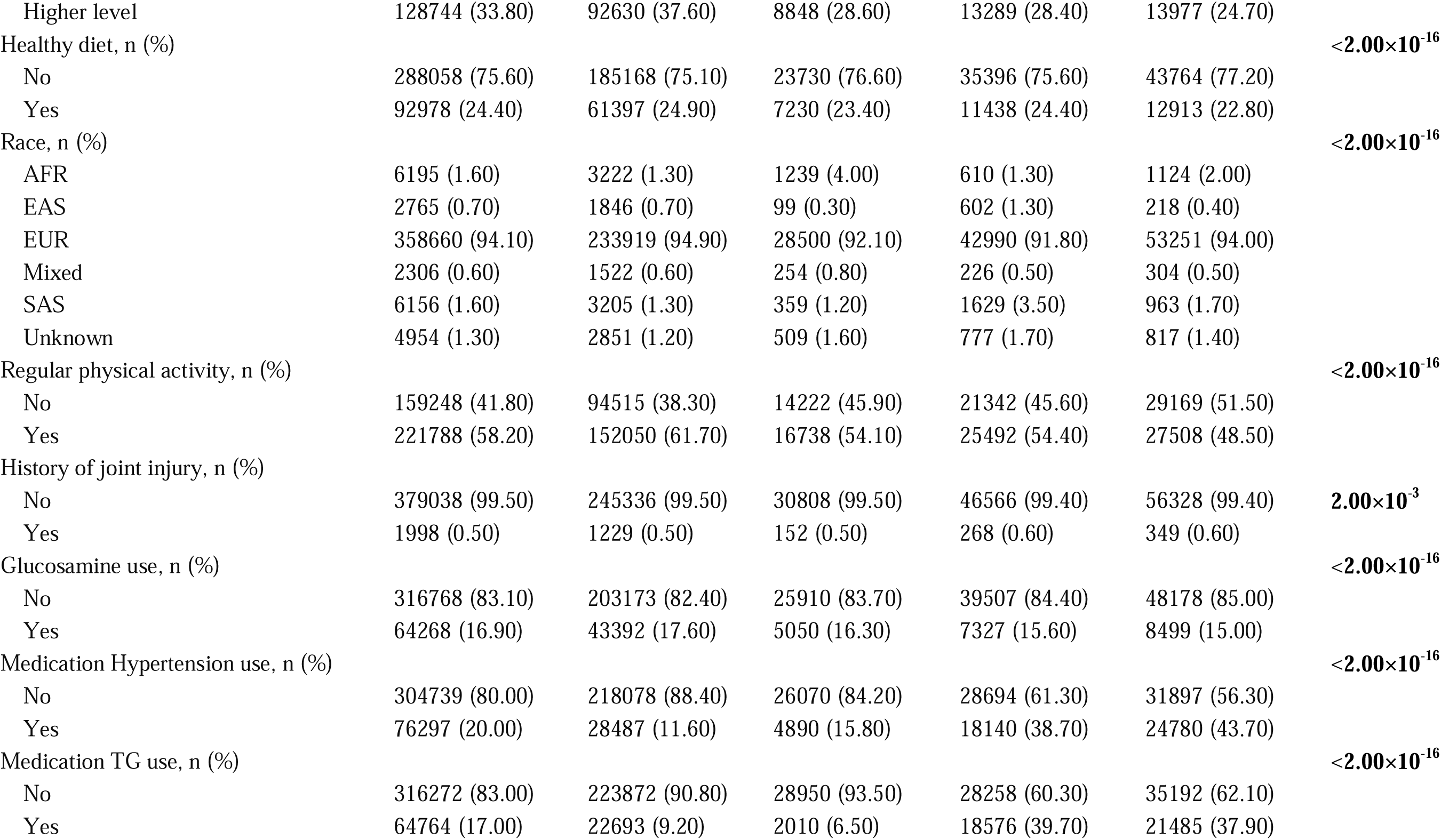
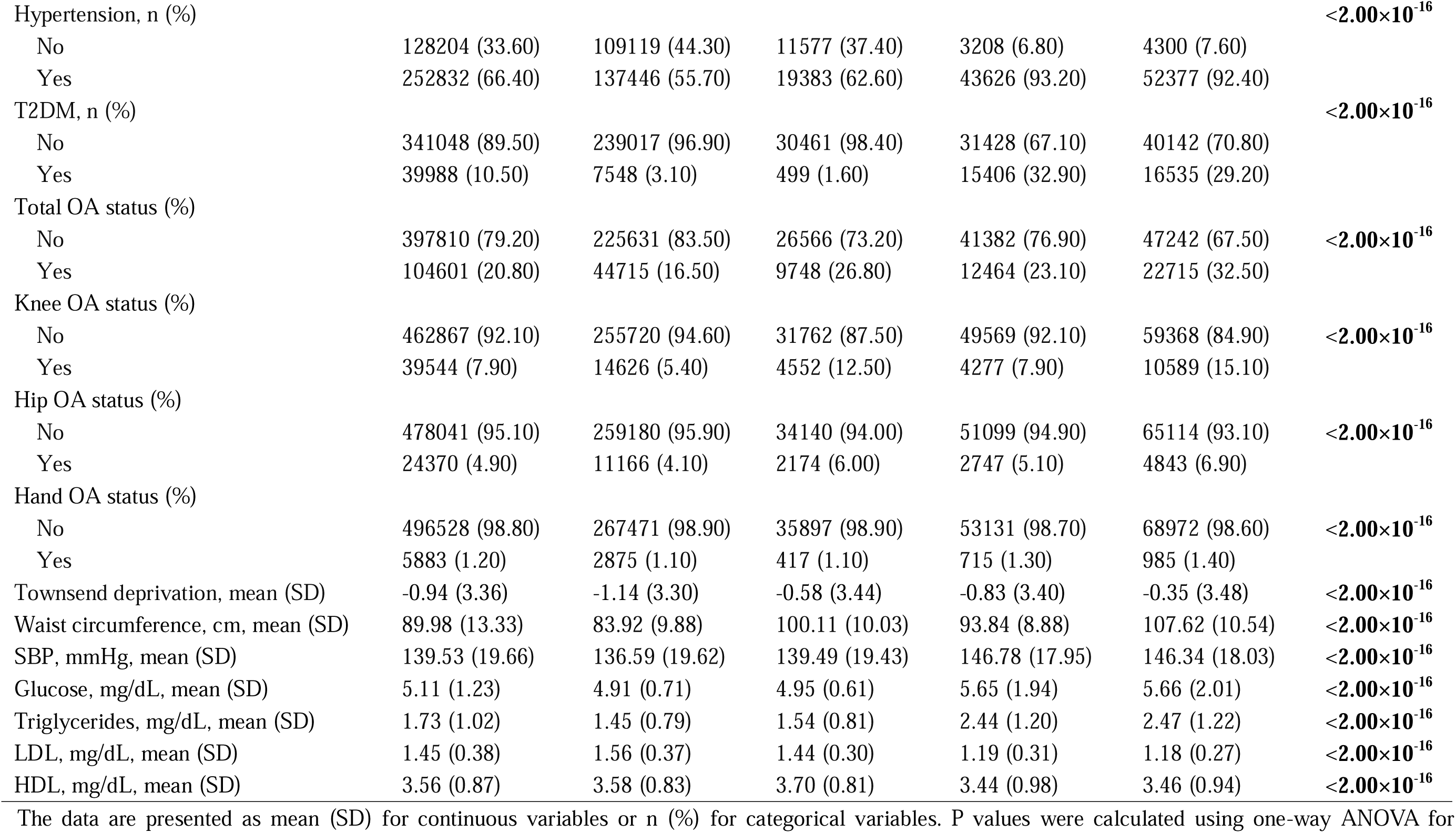
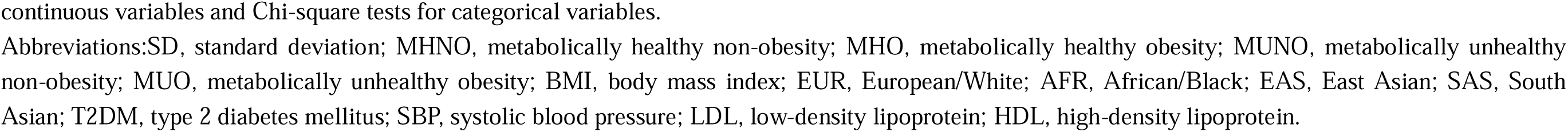
Baseline characteristics stratified by metabolic heterogeneity of obesity in the UK Biobank cohorts.

### Association between metabolic heterogeneity of obesity and incident OA

A total of 104,601 individuals (20.8%) were diagnosed with OA, including 39,544 (7.9%) with knee OA, 24,370 (4.9%) with hip OA, and 5,883 (1.2%) with hand OA (Table 1). The associations between metabolic heterogeneity of obesity and OA were examined on two models (crude model and fully adjusted model), constructed through Cox proportional hazards regression analysis. Both obesity and metabolically unhealthy status were consistently associated with a higher risk of OA across all models (Figure 2). Compared with MHNO individuals, the fully adjusted HRs for MUNO, MHO, and MUO were higher in weight-bearing joints. For total OA, HRs were 1.20 (1.17, 1.23), 1.72 (1.68, 1.77), and 1.87 (1.83, 1.91), respectively. For knee OA, HRs were 1.23 (1.18, 1.29), 2.42 (2.31, 2.53), and 2.56 (2.47, 2.66). For hip OA, HRs were 1.04 (0.99, 1.10), 1.51 (1.42, 1.61), and 1.49 (1.42, 1.56). In the non–weight-bearing joint (hand OA), HRs were 1.18 (1.06, 1.31), 1.08 (0.94, 1.24), and 1.13 (1.02, 1.26) (Figure 2).

**Figure 2.**
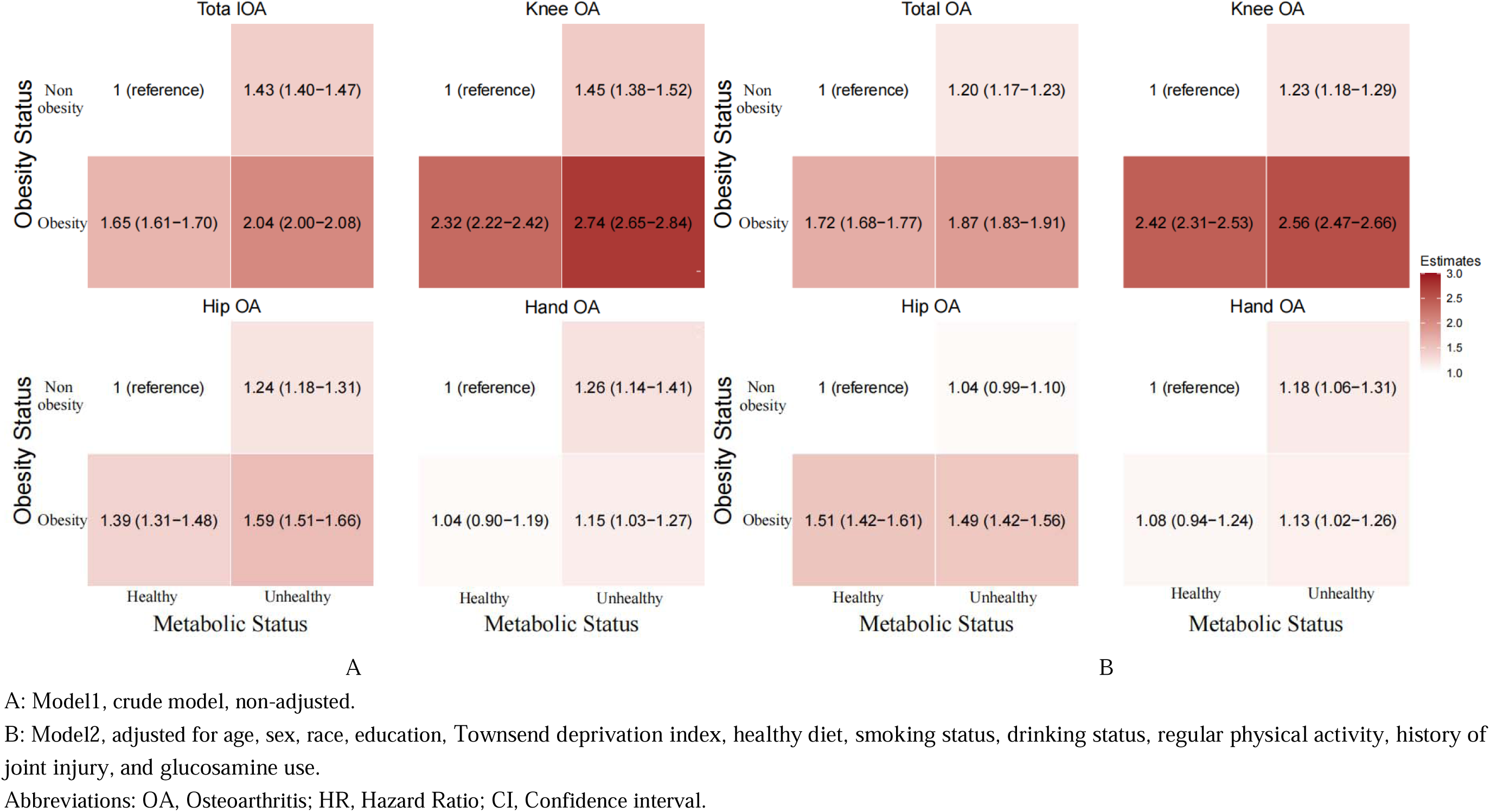
Cox regression analysis of the association between metabolic heterogeneity of obesity and the risk of incident OA.

The KM curves showed cumulative OA risk across metabolic heterogeneity of obesity groups over 12.4 years of follow-up, evaluated at 3-year intervals. Percentages denote cumulative incidence (Supplementary Figure 1). By 12 years, the MUO group had the highest cumulative incidence for total OA (28.82%), knee OA (10.49%), and hip OA (4.81%). For hand OA, the MUNO group had the highest cumulative incidence (0.91%), slightly exceeding MUO (0.87%). The MHNO group consistently had the lowest cumulative incidence across all sites: total OA (13.85%), knee OA (3.79%), hip OA (3.03%), and hand OA (0.73%). Differences between groups became more pronounced over time, particularly for total OA and knee OA (Supplementary Table 3).

### Mediation analysis of metabolic status in the relation between BMI and incident OA

The mediation analysis showed that metabolic factors played different roles in the relationship between BMI and incident OA at different sites (Supplementary Table 4). For total OA and knee OA, both the direct effects of BMI (HR 1.64 [1.60, 1.67] and 2.26 [2.18, 2.33], respectively; *P* < 0.05) and the indirect effects mediated by metabolic factors (HR 1.07 [1.06, 1.08] and 1.07 [1.05, 1.09]; *P* < 0.05) were significant, with proportions mediated of 15% and 11%, respectively. For hip OA, only the direct effect was significant (HR 1.47 [1.40, 1.53]; *P* < 0.05), with a non-significant indirect effect (HR 1.01 [0.99, 1.03]; *P* = 0.35). For hand OA, the total effect of BMI was not significant (HR 1.08 [0.99, 1.17]; *P* = 0.09), although a small but statistically significant indirect effect was observed (HR 1.06 [1.01, 1.11]; *P* < 0.05).

### Subgroup analysis

Consistent associations between metabolic heterogeneity of obesity and incident OA were observed across age and sex subgroups (Supplementary Figures 2–5). Significant interactions by age and sex were detected for total OA (*P*<0.001 for both) and knee OA (*P*<0.001 for sex; *P*=0.004 for age). For hip OA, the association was stronger in women (*P*=0.004), with no significant interaction by age (*P*=0.15). For hand OA, no significant interactions were identified in either subgroup.

### Sensitivity analysis

Sensitivity analyses were conducted by excluding participants with OA occurring within the first year and those occurring within the second year. After multivariable adjustment, metabolic heterogeneity of obesity remained a significant risk factor for incident OA at different sites (Supplementary Table 5-6).

## Discussion

In this study, we systematically examined the association between metabolic heterogeneity of obesity and the incidence of OA across multiple joint sites. We observed that metabolic factors had only a limited contribution to the risk of non–weight-bearing joints, such as hand OA. In contrast, for weight-bearing joints, particularly the knee, the combination of metabolic unhealthiness and obesity exerted a synergistic effect, leading to a substantially higher risk of OA. Furthermore, causal mediation analyses revealed that metabolic factors partially mediated the relationship between BMI and OA in weight-bearing joints, highlighting the complex interplay between adiposity and metabolic health in OA pathogenesis.

For non–weight-bearing joints such as hand OA, no significant associations were observed with BMI, and only a modest effect of metabolic factors, a finding that differs from some previous studies reporting stronger links between metabolic dysregulation and hand OA risk(18). This discrepancy may reflect the distinct pathogenesis of hand OA, in which aging, genetic predisposition, and local biochemical or inflammatory processes outweigh systemic metabolic influences(19, 20). These results underscore the need for future studies to integrate genomic, proteomic, and tissue-level analyses to better elucidate non-metabolic pathways in hand OA and to inform more targeted prevention strategies for non–weight-bearing joints.

In weight-bearing joints, elevated BMI promotes OA development primarily through increased mechanical stress, which alters the biomechanical environment, accelerates cartilage degeneration, and induces microtrauma and subchondral bone remodeling, leading to progressive cartilage breakdown over time(21, 22). As a result, these processes trigger a cascade of degenerative changes, including loss of cartilage integrity, increased subchondral bone density, and osteophyte formation(23). Beyond simple overload, excess body weight amplifies abnormal load distribution across the articular surface, particularly in the knee and hip, causing focal areas of high pressure that accelerate cartilage matrix degradation(24). Repetitive loading also induces microdamage in subchondral bone, triggering remodeling and sclerosis that alter joint mechanics and further stress the cartilage(25). Additionally, elevated BMI contributes to muscle weakness and joint malalignment, reducing shock absorption and exacerbating mechanical strain on weight-bearing joints(26). Given this strong mechanical component, maintaining a healthy weight and engaging in targeted physical activity may help reduce excessive joint loading and delay OA onset and progression in weight-bearing joints.

Our study found that metabolic factors play a crucial role in mediating the contribution of BMI to OA in weight-bearing joints. In these joints, visceral fat secretes pro-inflammatory cytokines like TNF-α, IL-6, and IL-1β, which trigger synovial inflammation and accelerate cartilage breakdown by increasing the production of MMPs and aggrecans(27, 28). Elevated leptin in obesity increases MMPs and inflammatory mediators in chondrocytes, accelerating cartilage destruction(29). Additionally, obesity acts as a key driver of metabolic syndrome, which subsequently promotes the production of reactive oxygen species (ROS)(30). These ROS directly damage chondrocytes and cartilage matrixes, while amplifying inflammation through pathways like NF-κB, creating a vicious cycle of cartilage degradation(31). Finally, metabolic dysfunction induced by obesity alters the joint microenvironment by triggering synovial inflammation and changing synovial fluid composition, impairing lubrication and exacerbating cartilage wear(32). As metabolic syndrome progresses, vascular dysfunction compromises nutrient delivery to avascular cartilage, further accelerating its degeneration.

The strengths of this study include its large population-based cohort, long-term follow-up, and the integration of metabolic heterogeneity into OA risk assessment(33). By moving beyond a BMI-only approach, our analysis provides a more nuanced understanding of OA risk and delivers a comprehensive joint-specific profile, highlighting distinct risk patterns between weight-bearing and non–weight-bearing joints. These findings emphasize the importance of addressing both weight control and metabolic health for effective OA prevention. In addition, incorporating routine metabolic status assessment into clinical practice is essential, as individuals with MUNO phenotypes are often overlooked due to their normal BMI. Regular metabolic screening can support early risk stratification and targeted interventions, offering an opportunity to slow or prevent OA progression.

This study also has limitations. First, the definition of metabolic status lacks uniformity across studies, potentially affecting the comparability of our results. To address this issue, we utilized the most common definition, incorporating five metabolic components from the metabolic syndrome criteria(9). Second, factors that could affect the incidence of OA, such as analgesics, anti-inflammatory drugs, and disease-modifying OA drugs(34, 35), were not considered as potential confounding factors. However, by excluding baseline OA participants, our study ensured that the analysis of transitions in obesity and metabolic health status on OA incidence remains robust and reliable for primary prevention. Third, although we adjusted for multiple covariates, residual confounding or unmeasured factors, such as diet, exercise, and genetic susceptibility, may still influence our findings.

In conclusion, this study highlights the importance of maintaining and promoting metabolic health, particularly in overweight/obese individuals, to reduce OA risk. Metabolic factors were identified as key mediators of the association between BMI and OA in weight-bearing joints, emphasizing the need for targeted strategies addressing both metabolic status and obesity.

## Data Availability

This research has been conducted using the UK Biobank resource under Application ID 67654. Data are available upon application to the UK Biobank (https://www.ukbiobank.ac.uk/), subject to their terms and approval process.

https://www.ukbiobank.ac.uk/

## Author contributions

HZ, HY and ZZ designed the study. HZ, HY, BZ, ZL managed and analyzed the data. HZ wrote the first draft of the article. All authors contributed to the data interpretation and report preparation, providing intellectual content to the manuscript, and granting approval for submission.

## Data availability

The dataset supporting the conclusions of this article is available in the UK Biobank repository in https://www.ukbiobank.ac.uk/ (Application ID: 67654).

## Ethical approval

UK Biobank has received ethical approval from the UK National Health Service’s National Research Ethics Service (16/NW/0274).

## Acknowledgments

This study makes use of data from UK Biobank (Project ID: 67654) and we thank the UK Biobank participants and the UK Biobank team for generating an important research resource.

