## Supplementary Tables and Figures for "Associations between metabolic heterogeneity of obesity and osteoarthritis: A prospective cohort study"

###### Supplementary Table 1. Information about exposures, outcomes, and covariates.

|  | **UK Biobank** | |
| --- | --- | --- |
|  | **Code** | **Description** |
| **Exposures** | | |
| **Metabolic heterogeneity phenotypes** |  | MHNO (Metabolically Healthy Non-Obesity)  MHO (Metabolically Healthy Obesity)  MUNO (Metabolically Unhealthy Non-Obesity)  MUO (Metabolically Unhealthy Obesity) |
| **Outcomes** | | |
| **All OA** | ICD-10: MI5  M16  M17  M18  M19  M471  M472  M478  M479  M480  ICD-9: 7150  7152  7153  7158  7159 | Primary generalized (osteo)arthritis  Coxarthrosis (arthrosis of hip)  Gonarthrosis (arthrosis of knee)  Arthrosis of first carpometacarpal joint  Other arthrosis  Other spondylosis with myelopathy  Other spondylosis with radiculopathy  Other spondylosis  Spondylosis, unspecified  Spinal stenosis  Localised, primary osteoarthrosis and allied disorders  Localised, secondary osteoarthritis and allied disorders  Unspecified localised osteoarthritis and allied disorders  Osteoarthrosis/allied dis. more than one site but not generalized   Osteoarthrosis/allied dis. not spec. whether gen. or loc |
| **Hip OA** | ICD-10: M160  M161  M169  ICD-9:71535  71515 | Primary coxarthrosis, bilateral  Other primary coxarthrosis  Coxarthrosis, unspecified  Unspec. localised osteoarthrosis/allied dis. (pelvic region and thigh)  Localised prim. osteoarthrosis/allied dis. (pelvic region and thigh) |
| **Knee OA** | ICD-10: M170  M171  M179  ICD-9: 71536  71516 | Primary gonarthrosis, bilateral  Other primary gonarthrosis  Gonarthrosis, unspecified  Unspec. localised osteoarthrosis/allied dis. (lower leg)  Localised, primary ostcourthrosis and allied disorders (lower leg) |
| **Hand OA** | ICD10: M151  M152  M1 54  M180  M181  M189  M1904  M1994  ICD9: 71514  71534 | Heberden's nodes (with arthropathy)  Bouchard's nodes (with arthropathy)  Erosive (osteoarthrosis)  Primary arthrosis of first carpometacarpal joints, bilateral  Other primary arthrosis of first carpometacarpal joint  Arthrosis of first carpometacarpal joint, unspecified  Primary arthrosis of other joints (Hand  Arthrosis, unspecified (Hand)  Localised, primary osteoarthrosis and allied disorders (hand)  Unspec. localised osteoarthrosis/ allied dis. (hand) |
| **Survival time** | 53  40000 | Survival time was calculated from the date of baseline assessment to the date of death. For participants without a death record, survival time was calculated to the last follow-up date (October 31, 2021). |
| **Covariates** | | |
| **Age at baseline** | 21022 | Age of the participant on the day they attended an Initial Assessment Centre, truncated to whole year. |
| **Sex** | 31 | Sex at baseline (male or female). |
| **Race** | 21000 | Ethnic background. |
| **Body mass index** | 23104 | Constructed from height and weight measured  during the initial Assessment Centre visit. |
| **Education** | 6138 | Qualifications of participant. |
| **Smoking status** | 20116 | The current/past smoking status of the participant. |
| **Alcohol intake** | 20117 | Alcohol drinker status. |
| **Regular physical activity** | 884  894  904  914 | Moderate physical activity  Vigorous physical activity  According to the International Physical Activity Questionnaire (IPAQ), exercise duration under 10 minutes per day is recorded as 0 minutes, and participants with over 960 minutes of daily PA are excluded as outliers. Moderate and vigorous time variables exceeding 240 minutes are truncated to 240 minutes. The total PA score ranges from 0 to 2, with 1 point assigned per met condition (MPA ≥ 55 min/week, VPA between 100-500 min/week), and PA is reclassified into low (0 points) and healthy (1-2 points) groups. |
| **Healthy diet** | Fruit:  1309  1319  Whole grains: 1438, 1448  1458, 1468  (Shell) fish:  1329  1339  Refined grains  1438, 1448  1458, 1468  Processed meats:  1349  3680 | Pieces fresh fruit/day (1 piece)  Pieces dried fruit/day (5 pieces)  Recommended intake≥ 3 servings/day  Whole meal/whole grain bread slices/week (1 slice/day)  Bran/oat/muesli cereal bowls/week (1 bowl/day)  Recommended intake≥ 3 servings/day  Oily fish/week (Once/week)  Non-oily fish/week (Once/week)  Recommended intake≥2 servings/week  White, brown, other bread slices/week (1 slice/day)  Biscuit, other cereals/week (1 bowl/day)  Recommended intake≤2 servings/day  Processed meat/week or daily (1 piece/day)  Age when last ate meat (0 pieces/day if indicated  having never eaten meat)  Recommended intake≤1 serving/week  A healthy diet was defined as an adequate intake of at least 3 of 5 food groups recommended |
| **Glucosamine use** | 6179 | Mineral and other dietary supplements |
| **Townsend deprivation index at recruitment** | 22189 | Based on the preceding national census output areas. Each participant is assigned a score corresponding to the output area in which their postcode is located. |
| **History of joint injury** | 41270 | Having a diagnosis recorded in hospital admission electronic health records (ICD-10 code:S700,$710,S73,S760,S800,S810,S820,S83, S870). |

###### Supplementary Table 2. Definitions of metabolic status.

|  | **Low risk range** | |
| --- | --- | --- |
| **ATPIII definition** | **Female** | **Male** |
| Waist circumference | ≤88 cm | ≤102 cm |
| Blood pressure | SBP < 135 mmHg, DBP < 85 mmHg, not on antihypertensive medications | SBP < 135 mmHg, DBP < 85 mmHg, not on antihypertensive medications |
| Fasting plasma glucose | <6.11 mmol/L, without self-reported diabetes | <6.11 mmol/L, without self-reported diabetes |
| High-density lipoprotein | >1.30 mmol/L | >1.04 mmol/L |
| Triglyceride | <1.70 mmol/L, not on lipid-lowering medications | <1.70 mmol/L, not on lipid-lowering medications |

Metabolic health was defined as meeting at least three of the above criteria; meeting fewer than three was classified as metabolically unhealthy.

| Supplementary Table 3. Follow-Up cumulative risk of OA among groups of different metabolic heterogeneity of obesity levels. | | | | | |
| --- | --- | --- | --- | --- | --- |
| **Outcome** | **Group** | **Follow-up time**  **(Years)** | **Number at risk (n)** | **Number of events (n)** | **Cumulative incidence (%)** |
| Total OA | MHNO | 0 | 246565 | 0 | 0 |
|  |  | 3 | 240297 | 4731 | 1.97 |
|  |  | 6 | 231436 | 6407 | 4.74 |
|  |  | 9 | 220160 | 8237 | 8.48 |
|  |  | 12 | 154067 | 8271 | 13.85 |
|  | MHO | 0 | 30960 | 0 | 0 |
|  |  | 3 | 29705 | 1049 | 3.53 |
|  |  | 6 | 28069 | 1351 | 8.34 |
|  |  | 9 | 26045 | 1677 | 14.78 |
|  |  | 12 | 17511 | 1447 | 23.05 |
|  | MUNO | 0 | 46834 | 0 | 0 |
|  |  | 3 | 44929 | 1309 | 2.91 |
|  |  | 6 | 42377 | 1721 | 6.97 |
|  |  | 9 | 39233 | 2080 | 12.28 |
|  |  | 12 | 26593 | 2102 | 20.18 |
|  | MUO | 0 | 56677 | 0 | 0 |
|  |  | 3 | 53479 | 2541 | 4.75 |
|  |  | 6 | 49519 | 2975 | 10.76 |
|  |  | 9 | 45065 | 3283 | 18.04 |
|  |  | 12 | 29945 | 3227 | 28.82 |
| Knee OA | MHNO | 0 | 246565 | 0 | 0 |
|  |  | 3 | 243437 | 1535 | 0.63 |
|  |  | 6 | 238804 | 2023 | 1.48 |
|  |  | 9 | 233087 | 2295 | 2.46 |
|  |  | 12 | 167426 | 2218 | 3.79 |
|  | MHO | 0 | 30960 | 0 | 0 |
|  |  | 3 | 30285 | 464 | 1.53 |
|  |  | 6 | 29406 | 562 | 3.44 |
|  |  | 9 | 28310 | 672 | 5.82 |
|  |  | 12 | 19683 | 593 | 8.83 |
|  | MUNO | 0 | 46834 | 0 | 0 |
|  |  | 3 | 45814 | 402 | 0.88 |
|  |  | 6 | 44367 | 546 | 2.11 |
|  |  | 9 | 42525 | 599 | 3.52 |
|  |  | 12 | 29914 | 605 | 5.54 |
|  | MUO | 0 | 56677 | 0 | 0 |
|  |  | 3 | 54929 | 1057 | 1.92 |
|  |  | 6 | 52598 | 1239 | 4.28 |
|  |  | 9 | 49860 | 1362 | 7.01 |
|  |  | 12 | 34523 | 1202 | 10.49 |
| Hip OA | MHNO | 0 | 246565 | 0 | 0 |
|  |  | 3 | 244122 | 852 | 0.35 |
|  |  | 6 | 239998 | 1513 | 0.98 |
|  |  | 9 | 234505 | 2043 | 1.85 |
|  |  | 12 | 168633 | 1995 | 3.03 |
|  | MHO | 0 | 30960 | 0 | 0 |
|  |  | 3 | 30594 | 152 | 0.50 |
|  |  | 6 | 30000 | 275 | 1.41 |
|  |  | 9 | 29191 | 372 | 2.69 |
|  |  | 12 | 20533 | 314 | 4.22 |
|  | MUNO | 0 | 46834 | 0 | 0 |
|  |  | 3 | 45993 | 223 | 0.48 |
|  |  | 6 | 44732 | 349 | 1.27 |
|  |  | 9 | 43018 | 469 | 2.36 |
|  |  | 12 | 30385 | 440 | 3.8 |
|  | MUO | 0 | 56677 | 0 | 0 |
|  |  | 3 | 55569 | 412 | 0.74 |
|  |  | 6 | 53908 | 545 | 1.75 |
|  |  | 9 | 51820 | 667 | 3.04 |
|  |  | 12 | 36245 | 641 | 4.81 |
| Hand OA | MHNO | 0 | 246565 | 0 | 0 |
|  |  | 3 | 246311 | 254 | 0.1 |
|  |  | 6 | 245970 | 341 | 0.24 |
|  |  | 9 | 245480 | 490 | 0.44 |
|  |  | 12 | 181321 | 528 | 0.73 |
|  | MHO | 0 | 30960 | 0 | 0 |
|  |  | 3 | 30938 | 22 | 0.07 |
|  |  | 6 | 30890 | 48 | 0.23 |
|  |  | 9 | 30816 | 74 | 0.47 |
|  |  | 12 | 22362 | 65 | 0.76 |
|  | MUNO | 0 | 46834 | 0 | 0 |
|  |  | 3 | 46767 | 67 | 0.14 |
|  |  | 6 | 46680 | 87 | 0.33 |
|  |  | 9 | 46577 | 103 | 0.55 |
|  |  | 12 | 34577 | 124 | 0.91 |
|  | MUO | 0 | 56677 | 0 | 0 |
|  |  | 3 | 56614 | 63 | 0.11 |
|  |  | 6 | 56513 | 101 | 0.29 |
|  |  | 9 | 56384 | 129 | 0.52 |
|  |  | 12 | 41465 | 144 | 0.87 |

Cumulative incidence values were derived from Kaplan–Meier survival analyses at 0, 3, 6, 9, and 12 years of follow-up.

Abbreviations: OA, osteoarthritis; MHNO, metabolically healthy non-obesity; MHO, metabolically healthy obesity; MUNO, metabolically unhealthy non-obesity; MUO, metabolically unhealthy obesity.

**Supplementary Table 4.** **Mediation analysis of metabolic factors in the association between BMI and incident OA.**

| **Outcome** | **Direct Effect**  **(HR [95% CI])** | ***P* value (Direct)** | **Indirect Effect (HR [95% CI])** | ***P* value (Indirect)** | **Total Effect (HR [95% CI])** | ***P* value (Total)** | **Proportion Mediated (pm [95% CI])** | ***P* value (pm)** |
| --- | --- | --- | --- | --- | --- | --- | --- | --- |
| Total OA | 1.64 [1.60, 1.67] | < 0.05 | 1.07 [1.06, 1.08] | < 0.05 | 1.76 [1.72, 1.79] | < 0.05 | 0.15 [0.13, 0.18] | < 0.05 |
| Knee OA | 2.26 [2.18, 2.33] | < 0.05 | 1.07 [1.05, 1.09] | < 0.05 | 2.41 [2.34, 2.49] | < 0.05 | 0.11 [0.08, 0.14] | < 0.05 |
| Hip OA | 1.47 [1.40, 1.53] | < 0.05 | 1.01 [0.99, 1.03] | 0.35 | 1.49 [1.43, 1.54] | < 0.05 | 0.03 [-0.03, 0.10] | 0.35 |
| Hand OA | 1.02 [0.91, 1.12] | 0.73 | 1.06 [1.01, 1.11] | < 0.05 | 1.08 [0.99, 1.17] | 0.09 | 0.77 [-2.67, 6.51] | 0.10 |

Proportion mediated (pm) represents the proportion of the total effect of BMI on OA mediated by metabolic factors. Non-significant mediation is indicated where 95% CIs cross zero.

Abbreviations: HR, Hazard ratio; CI, Confidence interval; OA, Osteoarthritis; BMI, Body mass index.

###### Supplementary Table 5. Association of metabolic heterogeneity of obesity with OA risk after excluding those occurring OA within the first year

|  |  | **Number (n, %)** | **Model 1** | | **Model 2** | |
| --- | --- | --- | --- | --- | --- | --- |
|  |  |  | **HR (95% CI)** | ***P* value** | **HR (95% CI)** | ***P* value** |
| **OA site** | **Metabolic heterogeneity of obesity** |  |  |  |  |  |
| Total OA | MHNO | 245295 (64.82%) | 1.00 (reference) |  | 1.00 (reference) |  |
|  | MHO | 30668 (8.10%) | 1.65 (1.60, 1.69) | <**2.00×10^-16^** | 1.72 (1.67, 1.77) | <**2.00×10^-16^** |
|  | MUNO | 46489 (12.28%) | 1.44 (1.40, 1.47) | <**2.00×10^-16^** | 1.20 (1.17, 1.23) | <**2.00×10^-16^** |
|  | MUO | 55959 (14.80%) | 2.02 (1.98, 2.06) | <**2.00×10^-16^** | 1.86 (1.82, 1.90) | <**2.00×10^-16^** |
| Knee OA | MHNO | 245295 (64.82%) | 1.00 (reference) |  | 1.00 (reference) |  |
|  | MHO | 30668 (8.10%) | 2.31 (2.20, 2.42) | <**2.00×10^-16^** | 2.41 (2.30, 2.53) | <**2.00×10^-16^** |
|  | MUNO | 46489 (12.28%) | 1.46 (1.39, 1.53) | <**2.00×10^-16^** | 1.25 (1.19, 1.31) | <**2.00×10^-16^** |
|  | MUO | 55959 (14.80%) | 2.72 (2.62, 2.82) | <**2.00×10^-16^** | 2.55 (2.46, 2.65) | <**2.00×10^-16^** |
| Hip OA | MHNO | 245295 (64.82%) | 1.00 (reference) |  | 1.00 (reference) |  |
|  | MHO | 30668 (8.10%) | 1.38 (1.30, 1.47) | <**2.00×10^-16^** | 1.50 (1.41, 1.59) | <**2.00×10^-16^** |
|  | MUNO | 46489 (12.28%) | 1.23 (1.16, 1.30) | <**2.00×10^-16^** | 1.03 (0.98, 1.09) | 0.27 |
|  | MUO | 55959 (14.80%) | 1.55 (1.48, 1.63) | <**2.00×10^-16^** | 1.46 (1.39, 1.54) | <**2.00×10^-16^** |
| Hand OA | MHNO | 245295 (64.82%) | 1.00 (reference) |  | 1.00 (reference) |  |
|  | MHO | 30668 (8.10%) | 1.08 (0.94, 1.24) | 0.29 | 1.12 (0.98, 1.30) | 0.11 |
|  | MUNO | 46489 (12.28%) | 1.26 (1.12, 1.40) | <**2.00×10^-16^** | 1.18 (1.06, 1.32) | **4.00×10^-3^** |
|  | MUO | 55959 (14.80%) | 1.14 (1.03, 1.27) | **1.40×10^-2^** | 1.14 (1.02, 1.27) | **1.80×10^-2^** |

Model 1: Crude model, unadjusted.
Model 2: Adjusted for age, sex, race, education, Townsend deprivation index, healthy diet, smoking status, alcohol drinking status, regular physical activity, history of joint injury, and glucosamine use.

Abbreviations: HR, hazard ratio; CI, confidence interval; OA, osteoarthritis; MHNO, metabolically healthy non-obesity; MHO, metabolically healthy obesity; MUNO, metabolically unhealthy non-obesity; MUO, metabolically unhealthy obesity.

###### Supplementary Table 6. Association of metabolic heterogeneity of obesity with OA risk after excluding those occurring OA within the second year

|  |  | **Number (n, %)** | **Model 1** | | **Model 2** | |
| --- | --- | --- | --- | --- | --- | --- |
|  |  |  | **HR (95% CI)** | ***P* value** | **HR (95% CI)** | ***P* value** |
| **OA site** | **Metabolic heterogeneity of obesity** |  |  |  |  |  |
| Total OA | MHNO | 243587 (65.11%) | 1.00 (reference) |  | 1.00 (reference) |  |
|  | MHO | 30316 (8.11%) | 1.65 (1.60, 1.70) | <**2.00×10^-16^** | 1.72 (1.67, 1.77) | <**2.00×10^-16^** |
|  | MUNO | 45995 (12.30%) | 1.43 (1.39, 1.47) | <**2.00×10^-16^** | 1.20 (1.17, 1.23) | <**2.00×10^-16^** |
|  | MUO | 55062 (14.72%) | 2.00 (1.96, 2.04) | <**2.00×10^-16^** | 1.84 (1.80, 1.88) | <**2.00×10^-16^** |
| Knee OA | MHNO | 243587 (65.11%) | 1.00 (reference) |  | 1.00 (reference) |  |
|  | MHO | 30316 (8.11%) | 2.32 (2.21, 2.43) | <**2.00×10^-16^** | 2.42 (2.30, 2.54) | <**2.00×10^-16^** |
|  | MUNO | 45995 (12.30%) | 1.46 (1.38, 1.53) | <**2.00×10^-16^** | 1.24 (1.18, 1.31) | <**2.00×10^-16^** |
|  | MUO | 55062 (14.72%) | 2.68 (2.58, 2.79) | <**2.00×10^-16^** | 2.52 (2.42, 2.62) | <**2.00×10^-16^** |
| Hip OA | MHNO | 243587 (65.11%) | 1.00 (reference) |  | 1.00 (reference) |  |
|  | MHO | 30316 (8.11%) | 1.37 (1.29, 1.46) | <**2.00×10^-16^** | 1.49 (1.40, 1.59) | <**2.00×10^-16^** |
|  | MUNO | 45995 (12.30%) | 1.21 (1.14, 1.28) | <**2.00×10^-16^** | 1.02 (0.97, 1.09) | 0.41 |
|  | MUO | 55062 (14.72%) | 1.52 (1.45, 1.60) | <**2.00×10^-16^** | 1.44 (1.37, 1.52) | <**2.00×10^-16^** |
| Hand OA | MHNO | 243587 (65.11%) | 1.00 (reference) |  | 1.00 (reference) |  |
|  | MHO | 30316 (8.11%) | 1.09 (0.94, 1.26) | 0.24 | 1.14 (0.98, 1.32) | 0.08 |
|  | MUNO | 45995 (12.30%) | 1.26 (1.12, 1.42) | <**2.00×10^-16^** | 1.20 (1.06, 1.35) | **3.00×10^-3^** |
|  | MUO | 55062 (14.72%) | 1.15 (1.03, 1.28) | **2.00×10^-2^** | 1.15 (1.03, 1.29) | **1.60×10^-2^** |

Model 1: Crude model, unadjusted.
Model 2: Adjusted for age, sex, race, education, Townsend deprivation index, healthy diet, smoking status, alcohol drinking status, regular physical activity, history of joint injury, and glucosamine use.

Abbreviations: HR, hazard ratio; CI, confidence interval; OA, osteoarthritis; MHNO, metabolically healthy non-obesity; MHO, metabolically healthy obesity; MUNO, metabolically unhealthy non-obesity; MUO, metabolically unhealthy obesity.

###### Supplementary Figure 1. Kaplan–Meier plots for the cumulative risk of OA among groups of different metabolic heterogeneity of obesity levels.

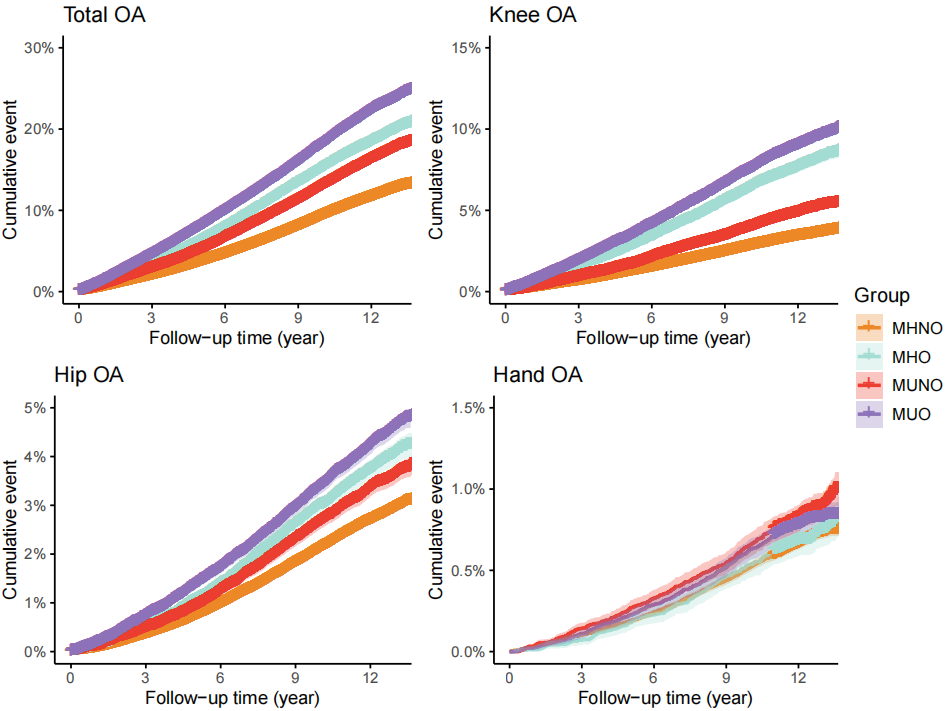

Abbreviations: OA, Osteoarthritis; MHNO, Metabolically healthy non-obesity; MHO, Metabolically healthy obesity; MUNO, Metabolically unhealthy non-obesity; MUO, Metabolically unhealthy obesity.

**Supplementary Figure 2. Subgroup analysis of the association between metabolic heterogeneity of obesity and incident total OA (Multivariate Cox Model).**

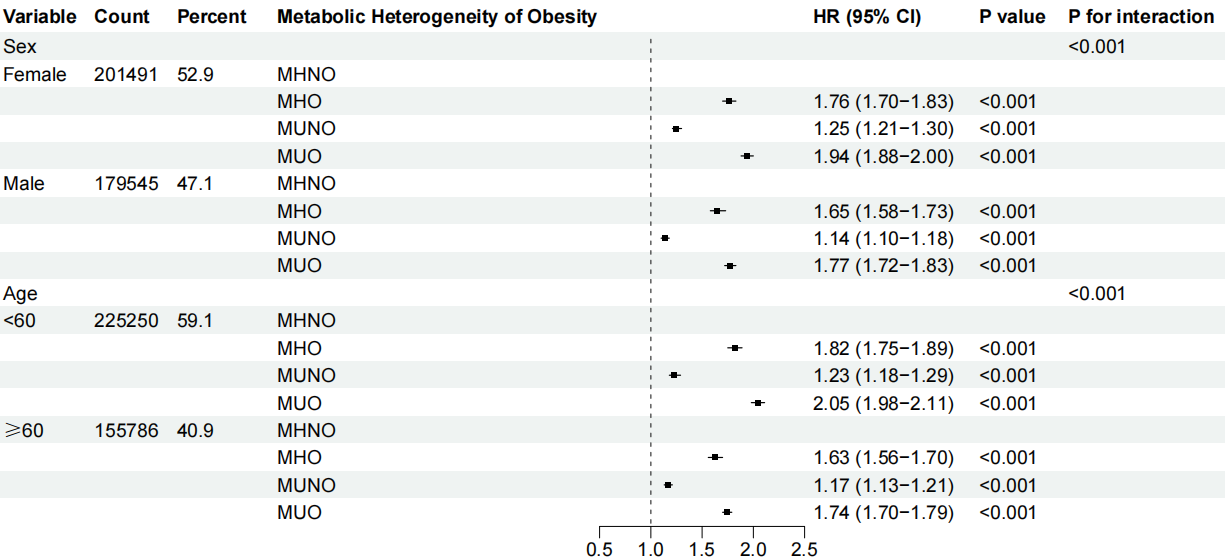

Cox regression adjusted for age, sex, race, education, Townsend deprivation index, healthy diet, smoking status, drinking status, regular physical activity, history of joint injury, and glucosamine use.

Abbreviations: HR, Hazard ratio; CI, Confidence interval; OA, Osteoarthritis; MHNO, Metabolically healthy non-obesity; MHO, Metabolically healthy obesity; MUNO, Metabolically unhealthy non-obesity; MUO, Metabolically unhealthy obesity..

**Supplementary Figure 3. Subgroup analysis of the association between metabolic heterogeneity of obesity and incident knee OA (Multivariate Cox Model).**

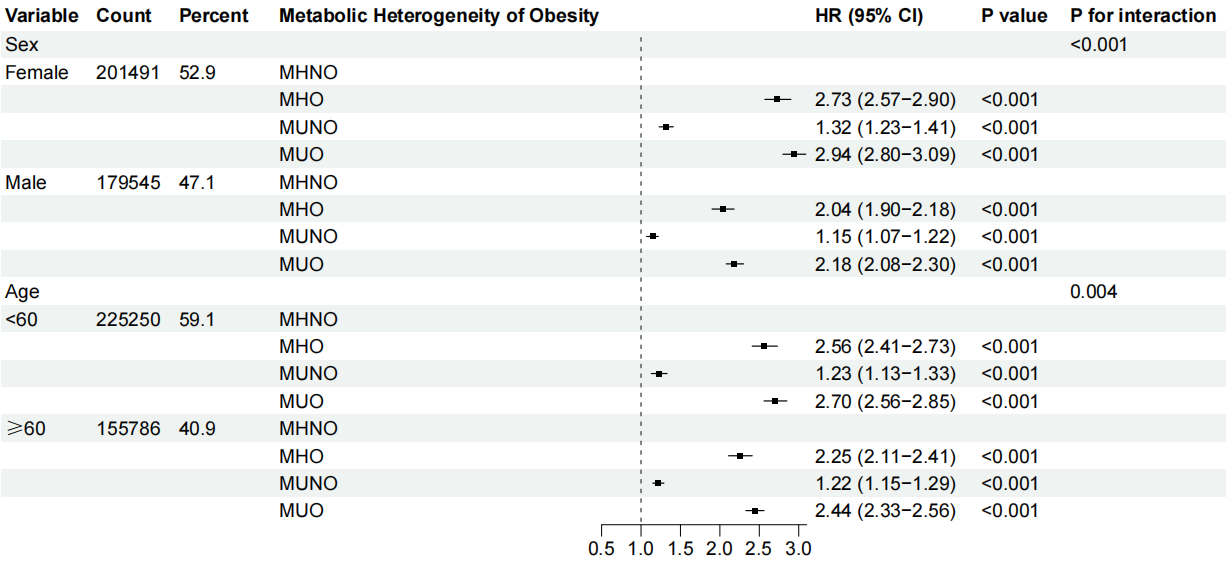

Cox regression adjusted for age, sex, race, education, Townsend deprivation index, healthy diet, smoking status, drinking status, regular physical activity, history of joint injury, and glucosamine use.

Abbreviations: HR, Hazard ratio; CI, Confidence interval; OA, Osteoarthritis; MHNO, Metabolically healthy non-obesity; MHO, Metabolically healthy obesity; MUNO, Metabolically unhealthy non-obesity; MUO, Metabolically unhealthy obesity.

**Supplementary Figure 4. Subgroup analysis of the association between metabolic heterogeneity of obesity and incident hip OA (Multivariate Cox Model).**

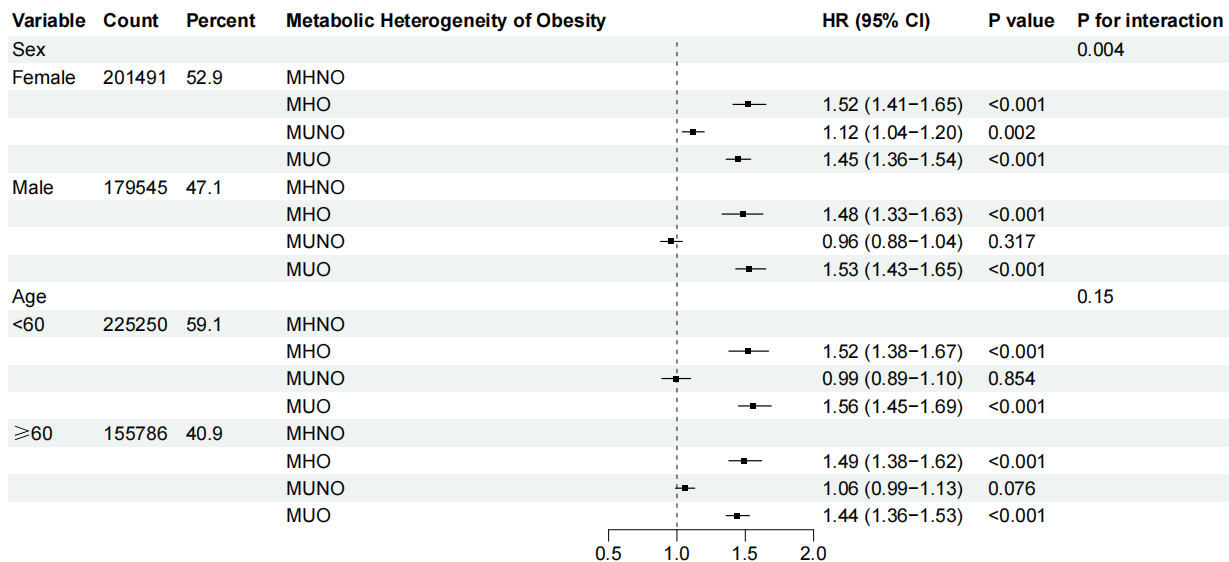

Cox regression adjusted for age, sex, race, education, Townsend deprivation index, healthy diet, smoking status, drinking status, regular physical activity, history of joint injury, and glucosamine use.

Abbreviations: HR, Hazard ratio; CI, Confidence interval; OA, Osteoarthritis; MHNO, Metabolically healthy non-obesity; MHO, Metabolically healthy obesity; MUNO, Metabolically unhealthy non-obesity; MUO, Metabolically unhealthy obesity.

**Supplementary Figure 5.** **Subgroup analysis of the association between metabolic heterogeneity of obesity and incident hand OA (Multivariate Cox Model).**

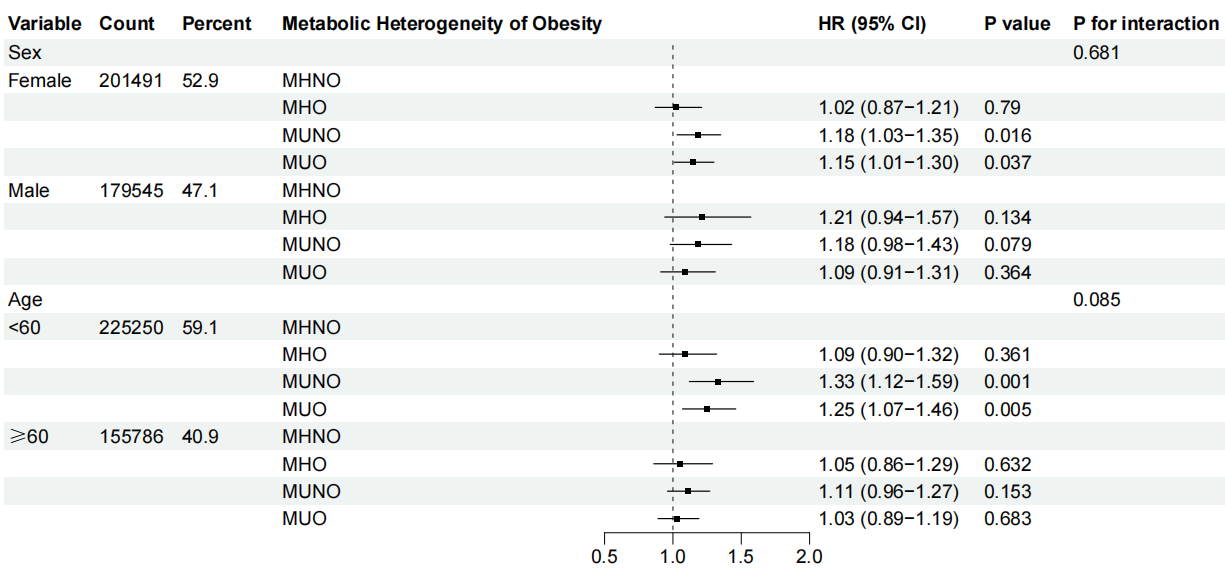

Cox regression adjusted for age, sex, race, education, Townsend deprivation index, healthy diet, smoking status, drinking status, regular physical activity, history of joint injury, and glucosamine use.

Abbreviations: HR, Hazard ratio; CI, Confidence interval; OA, Osteoarthritis; MHNO, Metabolically healthy non-obesity; MHO, Metabolically healthy obesity; MUNO, Metabolically unhealthy non-obesity; MUO, Metabolically unhealthy obesity.
